# Post Stroke Motor Recovery Genome Wide Association Study: *A Domain-Specific Approach*

**DOI:** 10.1101/2023.02.16.23286040

**Authors:** Chad M. Aldridge, Braun Robynne, Keith L. Keene, Fang-Chi Hsu, Michele M. Sale, Bradford B. Worrall

## Abstract

**Background:** In this genome wide association study (GWAS) we aimed to discover single nucleotide polymorphisms (SNPs) associated with motor recovery post-stroke.

**Methods:** We used the Vitamin Intervention for Stroke Prevention (VISP) dataset of 2,100 genotyped patients with non-disabling stroke. Of these, 488 patients had motor impairment at enrollment. Genotyped data underwent strict quality control and imputation. The GWAS utilized logistic regression models with generalized estimating equations (GEE) to leverage the repeated NIH Stroke Scale (NIHSS) motor score measurements spanning 6 time points over 24 months. The primary outcome was a decrease in the motor drift score of ≥ 1 vs. *<* 1 at each timepoint. Our model estimated the odds ratio of motor improvement for each SNP after adjusting for age, sex, race, days from stroke to visit, initial motor score, VISP treatment arm, and principal components.

**Results:** Although no associations reached genome-wide significance (p *<* 5 × 10^−8^), our analysis detected 115 suggestive associations (p *<* 5 × 10^−6^). Notably, we found multiple SNP clusters near genes with plausible neuronal repair biology mechanisms. The CLDN23 gene had the most convincing association which affects blood-brain barrier integrity, neurodevelopment, and immune cell transmigration.

**Conclusion:** We identified novel suggestive genetic associations with the first ever motor-specific post stroke recovery GWAS. The results seem to describe a distinct stroke recovery phenotype compared to prior genetic stroke outcome studies that use outcome measures, like the mRS. Replication and further mechanistic investigation are warranted. Additionally, this study demonstrated a proof-of-principle approach to optimize statistical efficiency with longitudinal datasets for genetic discovery.

## 1 Introduction

A reckoning is coming to the field of stroke recovery and genomics. The research, now merging at the intersection of these fields, faces three major challenges. First, a majority of the studies on stroke-related genes use a candidate gene approach [1], while there are only two genome-wide association study published to date [2, 3]. Current understanding of stroke recovery genetics is therefore limited to an extremely small portion of the genome, encompassing only 11 associated genes [1]. However the complex and time-varying biology of stroke recovery is likely to involve a much greater proportion of the genome. This suggests that study designs using genome-wide [4] and epigenome-wide [5] associations are well suited to discover novel recovery-associated genes and their variations. The second issue is that acute stroke treatment trials often collect blood samples useful for subsequent genetic studies. However, they tend to lack detailed measures of stroke recovery. Conversely, stroke recovery trials frequently collect these detailed and domain-specific outcome measures, but lack biospecimens for subsequent genetic analyses. The third challenge entails the issue that most studies on stroke recovery-related genes have defined their recovery phenotypes using global outcome measures that combine multiple domains of impairment (e.g. the modified Rankin Scale or total NIH Stroke Scale score) rather than using domain-specific measures (e.g. the Upper Extremity Fugl-Meyer for the motor domain) [6]. It remains unclear whether the phenotype-genotype associations observed using multi-domain measures differ from those observed using domain-specific measures of stroke recovery. For example variants of the BDNF (brain derived neurotrophic factor) gene have shown to predict poor stroke outcomes defined as the 90-day modified Rankin Scale (mRS) score ≤ 1 for ischemic stroke, or Glascow Outcome Scale score ≤ 3 for hemorrhagic stroke [7]. However, Cramer and colleagues [8] recently showed that BDNF variants were not associated with a domain-specific measure of arm motor function. This suggests that change in a multi-domain outcome measure may represent a different phenotype-genotype relationship than change in a domain-specific measure. The distinction is not trivial. As noted in the Stroke Recovery and Rehabilitation Roundtable [9] guidelines, “brain repair maps best onto fine-grained movement quality measures that are sensitive and specific.” In other words, using domain-specific measures of stroke recovery is better suited for studies that aim to discover genetic mechanisms of brain plasticity. Thus, genetic studies of stroke recovery using domain-specific measures are urgently needed. In an effort to address this need, Braun and colleagues [10] argued that changes in NIHSS subscores, which measure impairment in distinct neurological domains, can be considered as an efficient and clinically feasible means to obtain domain-specific measures of stroke recovery. They noted that the NIHSS motor impairment subscores are comparable to the Fugl-Meyer in terms of being specific to arm and leg motor function. They also have good inter-rater reliability (kappa 0.77-0.78). The present study is the first effort to define a phenotype-genotype association specific to post stroke motor-recovery using the change in NIHSS subscores.

## 2 Methods and Materials

### 2.1 Discovery Cohort

The Vitamin Intervention for Stroke Prevention trial (VISP) investigated the effect of vitamin supplementation dose on the risk of recurrent stroke with a randomized double-blinded design. The study enrolled patients who had a non-disabling ischemic stroke (mRS *<* 3) ≥ 72 hours prior to enrollment. Patients were randomized to a high dose or low dose vitamin supplementation arm if they were at least 75% compliant of taking a low dose supplementation packet for one month prior. All patients were reassessed every 3 months until a recurrent stroke event, but not longer than 2 years [11]. The trial successfully enrolled a total of 3680 randomized patients. This study was approved by the internal review boards (IRBs) of Wake Forest University School of Medicine, University of North Carolina at Chapel Hill School of Medicine as well as individual recruiting sites in accordance with the declaration of Helsinki. All patients provided written informed consent [12]. However, ten sites did not approve the genetic portion of the study resulting in 2,100 genotyped patients. We included only those that had a motor drift weakness of an arm or leg at the initial measurement of the NIH stroke scale at randomization. We excluded patients that had an incident recurrent stroke during the trial. This resulted in 488 participants in this GWAS.

### 2.2 Quality Control

The Center for Inherited Disease Research at Johns Hopkins University performed genotyping on the Illumina HumanOmni1-Quad-v1 array(Illumina, Inc.) The genotyped data underwent strict quality control measures that filtered out SNPs as follows: 1) missing call rate *>* 2%, 2) Mendelian errors in control trios, 3) deviation from Hardy-Weinburg equilibrium in controls, 4) discordant calls in duplicate samples, 5) sex differences in allele frequency or heterozygosity, 6) and minor allele frequency *<* 0.05 in line with previously published recommendations [13]. We further increased the number of SNP with genetic imputation via the TOPMed Imputation server [14, 15] which implements the Minimac Imputation procedure [16]. The TOPMed study [14] has a large cohort of 97,256 individuals with a diverse set of backgrounds which was preferred because of the sizeable proportion of non-European ancestry participants in the VISP genotyped cohort.

After filtering out imputed SNPs with poor imputation quality (*r*^2^ *<* 0.80) and MAFs *<* 0.05, the final count of SNPs came to 6,588,085.

### 2.3 Phenotyping

As suggested [10], we utilized the motor drift subscores of the NIHSS as a measurement of motor weakness. The NIHSS subscores 5A/5B and 6A/6B defined the degree of limb weakness for the upper and lower extremities also known as drift. The subscores rate limb weakness or drift on an ordinal scale from 0 to 5: 0 is no drift, 1 drift is present, 2 observed some effort against gravity, 3 shows no effort against gravity, 4 there is no movement, and 5 the limb is amputated. Motor improvement is defined as the decrease in the initial motor drift subscore of the weakest limb from enrollment to each follow up period. If patients had equally affected upper and lower limbs, we chose the upper limb. To maximize statistical power and model stability, we chose to dichotomize motor improvement as a decrease in initial motor drift by ≥ 1 versus *<* 1 for each follow up period.

### 2.4 Data Analysis Plan

We implemented a logistic regression model with generalized estimating equations (GEE) with the “gee” R package [17]. The GEE model allows the incorporation of repeated measurements of the motor drift subscore over the 2 year trial duration[18], which provides notable statistical power gains compared to the traditional case/control GWAS study design.

A priori we planned to adjust for age, sex, initial motor drift score, treatment arm, and population stratification via principle components. We calculated the top ten principle components utilizing the KING software [19] to account for population stratification in our cohort with genotyped SNPs after pruning. To determine which principle components to include in GWAS model, we used a backwards selection procedure optimizing the AIC with the “stepAIC” function from the MASS R package [20]. This approach allows for more efficient population stratification adjustment. In addition to the *a priori* covariates, time since stroke onset is an important covariate when modeling stroke recovery because of changing rates of recovery based on well defined time epochs(i.e. Acute, Early and Late Subacute, and Chronic)[21]. These epochs are tied to biological processes of inflammation and scarring early on into recovery with a transition to mainly endogenous plasticity in later stages. To account for this effect in the model, we added the covariate of time from stroke onset to time of motor drift measurement in days for each follow up period. Furthermore, we investigated the non-linear relationship of this covariate via binning the time from stroke onset to follow up period into quartiles. Figure 1 shows the mean estimated probability of motor drift improvement by each quartile. We decided to use a spline of time from onset to measurement with 1 knot at 250 days to better model the non-linear relationship and maintain clinical interpret-ability of the model for planned sensitivity analysis. Lastly, we considered possible loss to follow up effects. We planned to investigate which baseline characteristics predict missing motor drift scores. Any associated baseline characteristics would be added to the final GEE model with an exchangeable correlation structure as covariates.

**Figure 1:**
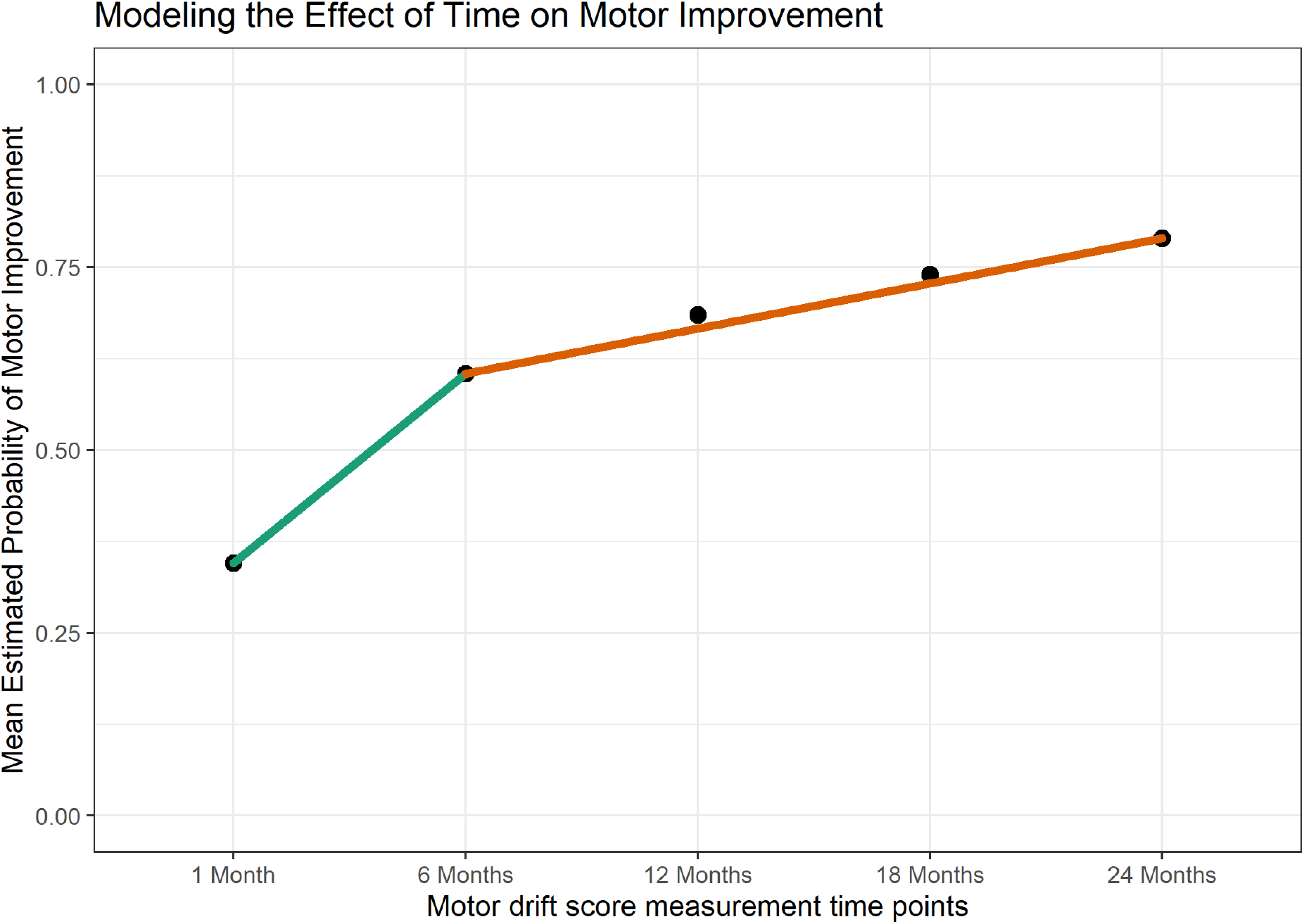
Shows the non-linear relationship of the mean probability of motor improvement for each follow up time point since study enrollment. The green and orange lines segments highlight the notable change in slopes from 1 to 6 month visits to 6 to 24 month visits. The difference in slope between the two time periods is similar to stroke rehab trials of the upper extremity.

### 2.5 Sensitivity Analysis

We performed two sensitivity analyses. First, we evaluated the interaction of time of stroke onset to follow-up period spline with each SNP that reached a p-value threshold of *p <* 5*x*10^−6^. We suspected that the effect of the SNP may change depending on the stroke recovery phase. Secondly, we observed a wide spread of time from stroke onset to VISP randomization (median 72 days; IQR 45.75 - 102 days). We generated an early versus late post-stroke enrollment variable defined as *<* the median (72 days) being early and ≥ the median as late enrollment, then estimate its interaction with each SNP as a separate sensitivity analysis.

### 2.6 Look-Up Analysis

We wished to investigate if the reported SNPs from the GISCOME GWAS study [2] on stroke functional recovery replicate with our post stroke motor recovery associated SNPs. The GISCOME study is the largest post stroke recovery GWAS by combined sample size (*n* = 6, 021) from 12 studies. Söderholm et al. defined good recovery as a mRS of ≤ 2 and a mRS of ≥ 3 signified poor recovery. We planned to compare our GWAS results with all SNPs with a p value *<* 5 *×* 10^−6^ from the GISCOME study. The p-values of the look-up analysis will receive a multiple comparison adjustment at a FDR of 10%. We chose the 10% rate because the look-up analysis has SNPs in linkage disequilibrium. The FDR algorithm assumes that each hypothesis test is independent from one another, which is violated when applied to SNPs within linkage disequilibrium. This violation biases the adjusted p-values to the null which makes a FDR of 5% exceedingly conservative.

## 3 Results

### 3.1 Demographics

The 488 patients provided 2,095 individual observations over the entire VISP study 2 year period from enrollment to months 1, 6, 12, 18 and 24. Patients had a median (IQR) 5 (4-5) number of motor drift assessments with a minimum of 1 to a maximum of 5. Twenty six point six percent of patients were lost to follow up by the 24 month visit. We found that sex and self-identified race were associated with loss to follow up. Males made up 76% patients that were lost to follow versus 59% (p*<* 0.001). Patients that identified as Black were more likely to be lost to follow up (30% vs 15%; p *<* 0.001). In contrast, self-identified White patients were less likely to be lost to follow up (62% versus 77%; p *<* 0.001). Table 1 shows the demographics of this cohort. As expected, most of the patients had worse arm weakness (67%) than leg weakness likely due to the VISP inclusion criteria of non-disability strokes defined by a mRS ≤ 3. It is well known that the higher mRS scores are biased toward lower extremity weakness and inability to walk compared to upper extremity weakness. Of note, the distribution of patient ancestry generally reflects the national U.S. population.

**Table 1:** Vitamin Intervention for Stroke Prevention (VISP) trial demographics. Information is presented as counts (percentages) or means (standard deviations).

| VISP Cohort (n=488) |  |
| --- | --- |
| **Treatment Arm** |  |
| High Dose | 225 (47%) |
| Low Dose | 251 (53%) |
| **Age in Years** |  |
| Mean (SD) | 66 ( ± 11) |
| **Sex** |  |
| Male | 310 (64%) |
| **Weakest Limb** |  |
| Arm | 333 (67%) |
| **Stroke Onset to Enrollment** |  |
| Days | 72.5 ( ± 31.2) |
| **Body Mass Index (BMI)** |  |
| N-Miss | 7 |
| Mean (SD) | 28.54 (6.47) |
| **Hypertension** |  |
| N-Miss | 1 |
| No | 114 (23%) |
| Yes | 383 (77%) |
| **Ever Smoker** |  |
| N-Miss | 1 |
| No | 173 (36%) |
| Yes | 314 (64%) |
| **Diabetes Mellitus Type II** |  |
| No | 331 (68%) |
| Yes | 157 (32%) |
| **Ancestry** |  |
| European | 347 (73%) |
| African | 92 (19%) |
| Other | 37 (8%) |

### 3.2 GWAS

#### 3.2.1 Primary Results

None of the SNPs reached genome-wide significance (p *<* 5 *×* 10^−8^). However, 115 SNPs reached suggestive associations with motor improvement (*p <* 5 *×* 10^−6^). Figure 2 plots the values of the odds ratio of motor improvement for each SNP. The calculated genomic control *λ* of the GWAS is 1.01, which suggests no genomic inflation. Therefore, we did not adjust the p-values.

**Figure 2:**
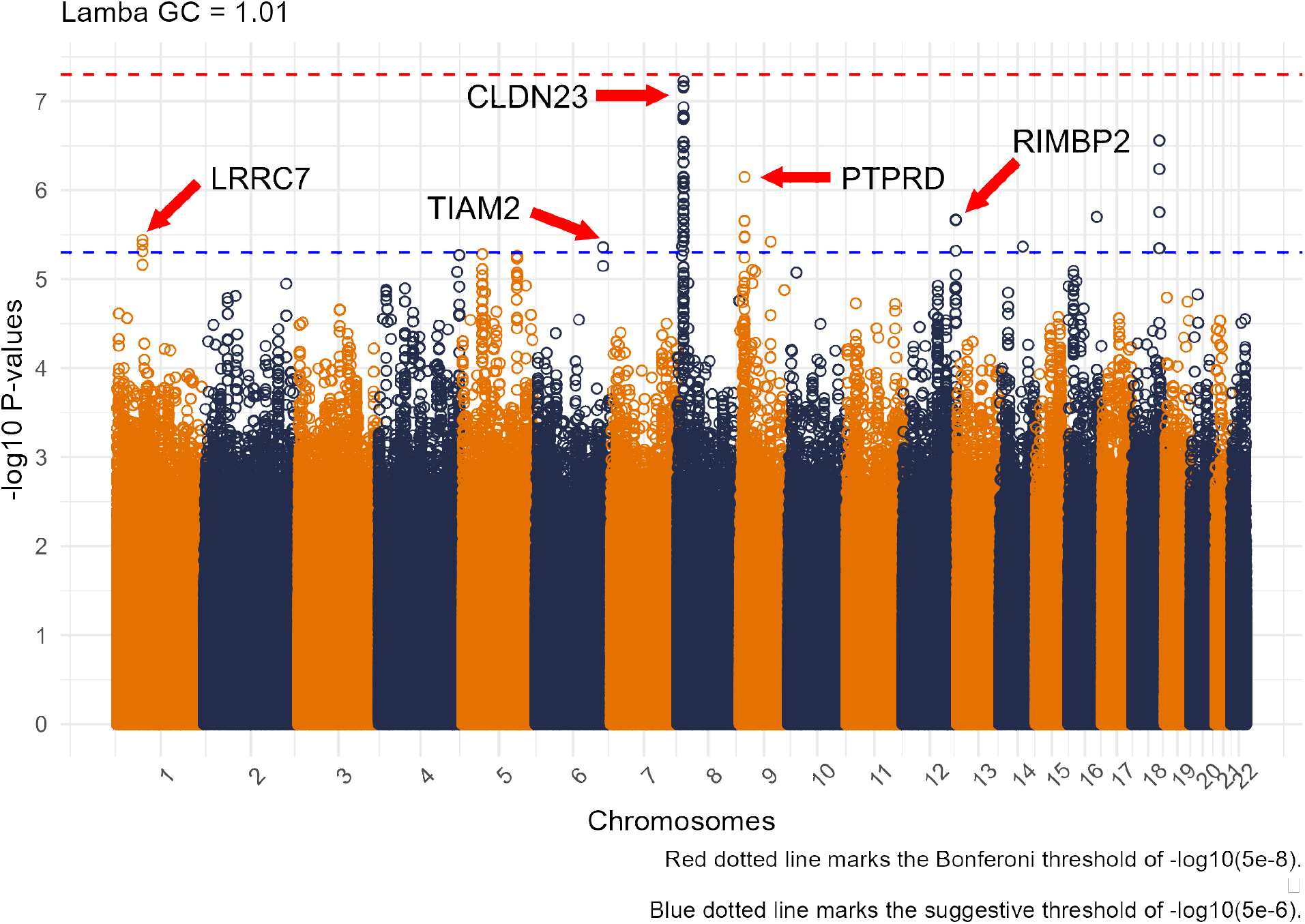
This Manhattan plot shows each SNP and its -log10(p value) associated with post stroke motor improvement. None of the SNPs reached genome-wide significant (above the red line). However, 115 SNPs had suggestive associations (above the blue line) with 2 right under the red line. The most convincing genetic loci is the large spike in chromosome 8; near the Claudin 23 gene. This gene affects blood brain barrier and immune cell transmigration.

The suggestive SNPs found themselves in chromosomes 1 (3), 6 (1), 8 (92), 9 (6), 12 (6), 14 (1), 16 (1), and 18 (5). The top two SNPs, rs12681936 and rs12680789, in chromosome 8 had the smallest p-values (5.96 *×* 10^−8^), which were just shy of genome-wide significance (*p <* 5 *×* 10^−8^). See Supplement Table 1 for a full list of all suggestive SNP associations with annotations from Ensembl.org’s variant effect predictor software [22]. Figure 2 shows a strong signal on chromosome 8. This locus is better visualized by the locus zoom plot in Figure 3 A. This locus is within *<* 0.1 megabases of the CLD23 gene.

**Figure 3:**
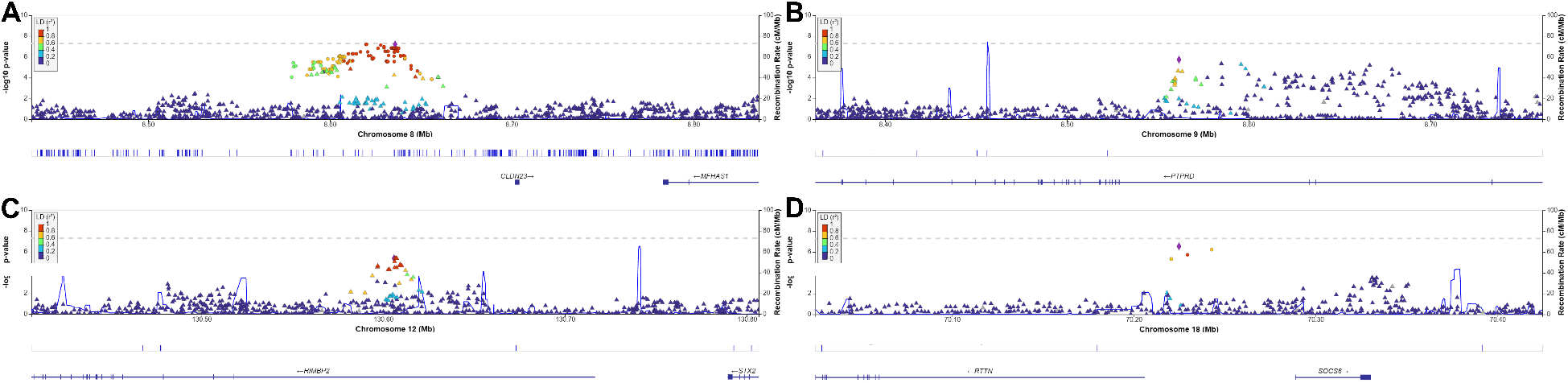
Panel plot of Locus Zoom figures (A-D) corresponding to genetic loci of interest. The colors refers to correlation of each SNP to the top SNP in each panel with red having an *r*^2^ ≥ 0.80. **A** Genetic locus near the CLDN23 gene on chromosome 8. **B** Genetic locus within the PTPRD gene on chromosome 9. **C** Genetic locus within the RIMBP2 gene on chromosome 12. **D** Genetic locus between the RTTN and SOCS6 genes on chromosome 18.

#### 3.2.2 Sensitivity Analysis

Sensitivity analysis of the interaction between the spline of stroke onset to motor drift measurement revealed that two SNPs had significant interactions at a FDR of 10%. They were rs113693489 in chromosome 6 and rs2967308 in chromosome 16. Supplement Table 2 contains all the interaction estimates and their q values. In general, SNP interactions with the first part of the spline (Days from stroke onset to measurement *<* 250) had a mean (*±* sd) Odds Ratio of 0.967 (*±* 1.33). The interactions with the second part of the spline (≥ 250 days) had a mean (*±* sd) Odds Ratio of 0.806 (*±* 1.33). Highlighting the chromosome 8 locus, Figure 4 shows the odds ratio point estimate and their 95% confidence intervals.

**Figure 4:**
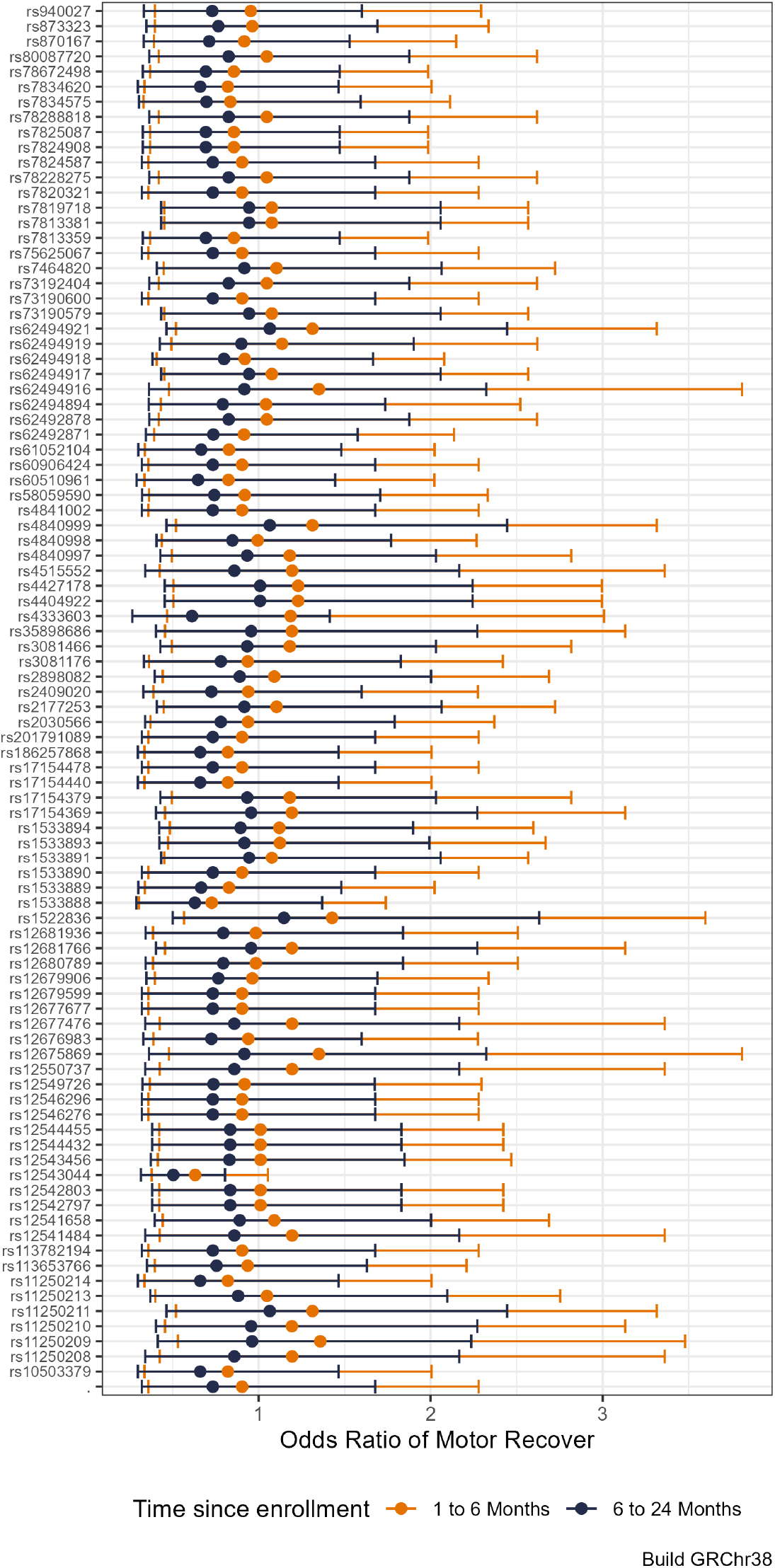
Shows the estimates and 95% confidence intervals of the interaction of the study timepoints 1 to 6 months versus 6 to 24 months with each suggestive SNP found in Chromosome 8. Interestingly, the interaction estimates of many SNPs seem to straddle one with SNP estimates at 1 to 6 months having a greater odds of motor recovery, while SNP estimates at 6 to 24 months having less.

The second sensitivity analysis focused on the interaction of the suggestive SNPs based on early versus later enrollment into the VISP trial from stroke onset. The SNPs’ p-values underwent a FDR adjustment of 10%. In contrast to the interaction analysis, all Early and Late enrollment values were significant; See Supplemental Table 3. Early enrollment interactions had a mean (*±* sd) Odds Ratio of 0.419 (*±* 1.68). Late enrollment interaction had a similar mean (*±* sd) Odds Ratio of 0.397 (*±* 1.70). Figure 5 exhibits the estimates for each suggestive SNP interaction with Early versus Late enrollment in the chromosome 8 locus. While our sensitivity analysis models show that each SNP interaction is an independent predictor of motor improvement, the 95% confidence intervals have large overlaps. The overlaps suggest that Odds Ratios for each SNP interaction do not differ from Early versus Late enrollment in the VISP trial from stroke onset.

**Figure 5:**
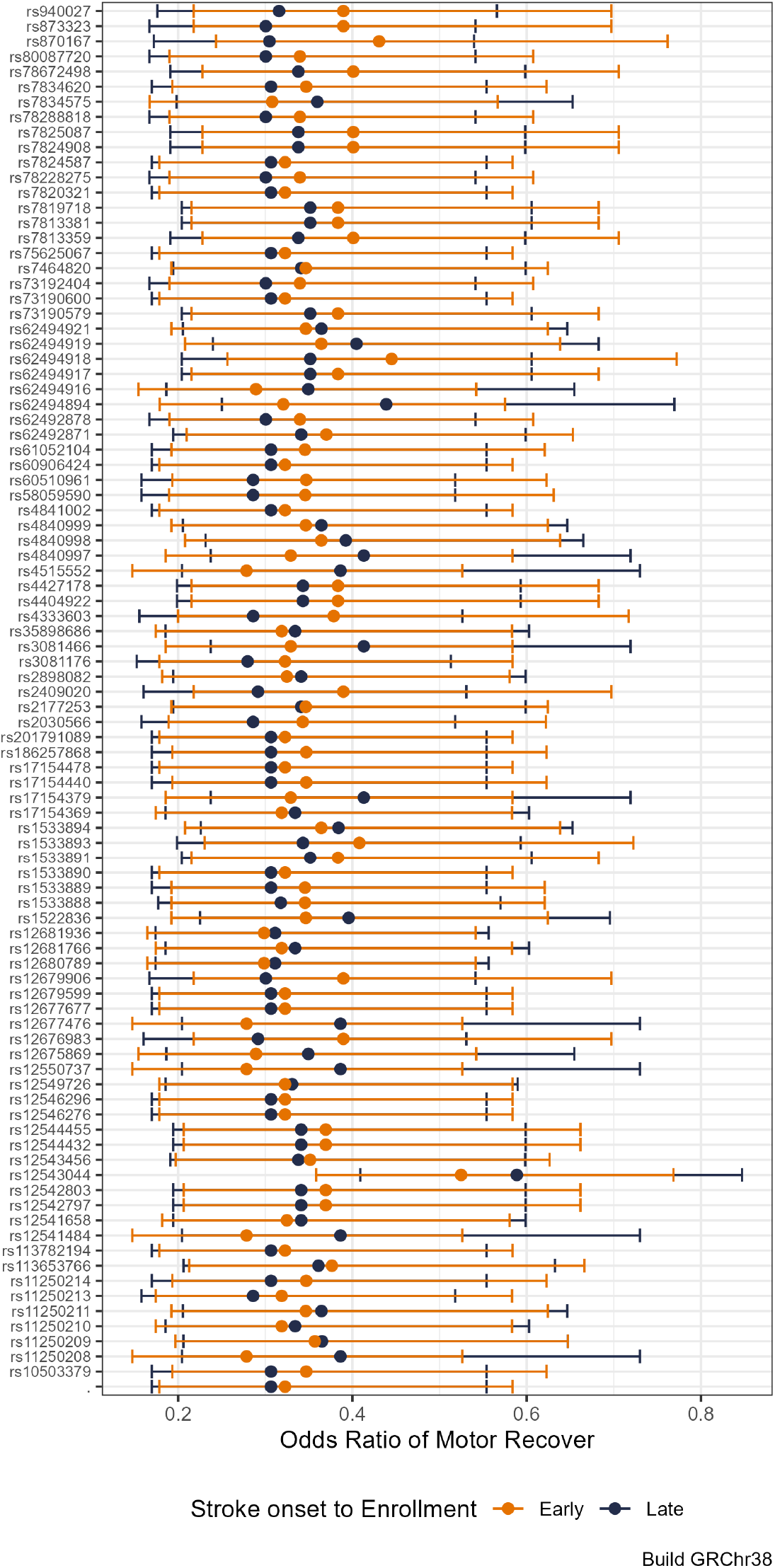
Shows odds ratios and 95% confidence intervals of motor drift score improvement from the interaction Early versus Late post-stroke enrollment by SNP in Chromosome 8. The odds ratios estimates for Early versus Late do not have a discernible pattern or consistency.

#### 3.2.3 Look-Up Analysis

Out of the 500 reported SNPs (p*<* 5 *×* 10^−6^) from the GISCOME study [2], only 414 were present in our analysis results. After applying a FDR of 10%, none of the look up SNPs from the GISCOME study reached significance.

## 4 Discussion

Our GWAS of post stroke motor recovery failed to show genome-wide significant associations. However, we found 115 novel suggestive SNPs linked to the odds of motor recovery over a two years in the first ever motor-specific post stroke recovery GWAS. These suggestive SNPs mapped to genomic loci connected to genes that are previously unknown as either candidate genes or ones from prior GWAS studies [2, 3]. Following from here, we discuss the genetic loci of interest associated with motor recovery individually.

### 4.1 Post Stroke Motor Recovery Genetic Loci

The Chromosome 8 locus’s apex, as seen in Figure 3 A, is *<* 0.1 megabases from the CLD23 gene. CLD23 (Claudin 23) is a protein encoding gene part of the Claudin gene family which are integral membrane proteins and components to tight junction strands[23]. CLDN23 has related pathways affecting the blood brain barrier and immune cell transmigration according to genecards.org’s pathway unification database (https://pathcards.genecards.org/). Additionally, CLDN23 variants are associated with blood cholesterol, triglyceride, and lipid measurements[24–26].

Unlike the chromosome 8 locus, the chromosome 9 locus finds itself within the PTPRD gene; part of the protein tyrosine phosphate (PTP) family. The PTPRD gene has a protein to protein interaction at the neuronal synapse located at the presynpatic terminal surface. It has related pathways of cell growth, differentiation, mitotic cycle, and oncogenic transformation[27, 28]. Interestingly, PTPRD has an association with glioblastoma [29].

Figure 3 C shows the chromosome 12 locus within in the RIMS-Binding Protein (RIMBP2) gene. As the name suggests, this gene produces a binding protein. The function of this protein is predicted to involve neuromuscular synpatic transmission. It is also highly expressed in brain tissue. Butola et al. in 2021 reported that the role of RIM-BP2 is to link voltaged-gated *Ca*^2+^ channels and release sites of synaptic vesicles[30]. They explain that RIMBP2 disruption leads to alterations in Cav2.1 channel topography at active zones. These active zones affect neurotransmitter release. The top SNP (rs73156962) of this locus has a direct biological interpretation (*p* = 0.034) in nucleus accumbens located in the basal ganglia, a highly dense interconnected neuronal tissue [31].

The chromosome 18 locus sits almost equally between two genes, RTTN and SOCS6, each within 0.1 megabases. See Figure 3 D. RTTN (Rotatin) encodes a large protein without a known specific function. However, knockout mice models result in neural tube defects [32]. In humans, RTTN pathological variants lead to microcephaly and polymicrogyria with seizures [33, 34]. Even though RTTN is linked to neurological structure and disorder in humans, there remains a notable lack of published literature on this gene and its biological mechanisms. However, SOCS6 (Supressor of Cytokine Signalling 6) is part of the supressor cytokine signalling protein family which plays a key role in inflammation regulation and insulin signalling in human brain tissue, especially brain tissue affected by a neuro-degenerative disease [35].

### 4.2 Literature Comparison

Söderholm et al. [2] performed the largest GWAS of stroke functional recovery to date. Their analysis consisted of 12 studies which totaled 6,021 patients. They defined functional stroke recovery as the obtainment of a mRS score ≤ 2 as “Good Recovery” versus ≥ 3 as “Poor Recovery” in their case/control approach. The study found only 1 SNP (rs1842681) significant at the genome-wide level (*p <* 5 *×* 10^−8^) located in the LOC105372028 gene. The LOC105372028 gene has no known biological function. They also found 33 suggestive SNPs among 12 different loci. When they utilized the mRS scores as a ordinal response instead of a binary one, the number of suggestive SNPs increased to 75 spread over 17 distinct loci without an increase in genome-wide significant SNPs.

When comparing Söderholm et al.’s study with ours, there are two notable distinctions. First, the set of associated genes of each study are unique. None of our associated genes replicated with theirs. This fact is intriguing because the unique set of genes from each study may be due to the functional recovery measures utilized. Our GWAS analysis used the motor drift scores from the NIHSS as a specific motor behavior marker. Plus, most of the patients in our discovery cohort had the greatest weakness in the upper extremity instead of the lower which suggests that motor drift score changes may not correlate with changes in the mRS. Unlike the motor drift score, the mRS encompasses multiple phenotypic domains like cognition, motor strength, balance, and mortality. The mRS measure has the probability of associating with genes that have general or systemic biological effects as well as include genes expressed in other tissues that interact with brain tissue like cardiovascular and lymphatic tissues.

The second distinction is the clear difference in the number of patients in each study, ours *n* = 488 and Söderholm et al. *n* = 6, 201. We capitalized on the repeated motor drift scores measurements. The logistic regression model with GEE greatly enhanced the statistical efficiency to find SNPs of interest associated with post-stroke motor recovery. In fact this study’s had about one-tenth the minimum recommended sample size for GWAS studies [36, 37]. Thus, our analysis is a proof-of-principle that longitudinal observational studies can be a strong design for future stroke recovery genomic studies.

To note, the genetic loci of Söderholm et al. and ours, did not include well published candidate stroke recovery genes of APOE, BDNF or COX-2 [1, 38–41]. This has particular interest since one would imagine that at least one of these genes would present themselves in either our results or those of Söderholm et al. Even more so in line with Söderholm et al. because of the use of the mRS as a recovery measure like previous candidate gene studies. One possible explanation is that GWAS studies remain too under-powered to detect the effect size of known candidate genes. The effect sizes of the candidate genes may be smaller than anticipated. To address the issue of being under-powered, the stroke recovery community needs more genetic data linked to specific stroke recovery phenotypes of interest or at least capitalize on observational stroke outcome studies with longitudinal designs overlaid on relevant recovery milestones.

### 4.3 Limitations

Unfortunately, our study does not have a replication cohort, despite searching internationally for other cohorts with NIHSS subscores and genetic data. For example, the National Institute of Neurological Disorders and Stroke (NINDS) archived clinical research database has 22 publicly available stroke study datasets, but none of them have NIHSS subscores and genetic data. However, we performed a look-up analysis based on Söderholm et al.’s findings [2] as a reasonable surrogate replication cohort. Another limitation of study is related to the VISP enrollment criteria. Patients enrolled into the VISP trial must have had a stroke due to atheroembolic mechanisms. Potential patients were excluded if their stroke was the result of a cardioembolic source. It is possible that our discovery cohort of patients had more small vessel strokes compared to large artery and other stroke types. The biological mechanisms deployed and their effect on stroke recovery may differ among these subtypes, especially since large artery and cardioembolic stroke tend to have larger stroke lesion volumes than small vessel strokes. Small vessel stroke may relate more to chronic inflammatory or hypertension exposures which may explain CLDN23 as the most promising finding. Unfortunately, the VISP trial did not collect stroke subtype data like the TOAST criteria [42]. We are unable to investigate how the genetic associations may differ among stroke subtypes.

## 5 Conclusion

We demonstrated the first ever use of repeated measurements and a domain-specific phenotype in a stroke recovery GWAS. This resulted in the discovery of new genes associations. As a proof-of-principle, this GWAS repurposed the NIHSS in a rich stroke clinical trial data set in line with the recommendations from Braun et al. [10] and the Stroke Recovery and Rehabilitation Roundtable [9]. This study’s approach may have a great impact on future genetic stroke study design. Longitudinal design allows one to investigate if the SNP effect is associated with changes over time, which we believe is critical in stroke recovery genetics research.

## Data Availability

All data produced in the present work are contained in the manuscript

## 6 Author Contributions

CA, RB, KK, FH, BW contributed to the conception and design of the study. MS and BW were instrumental in the acquisition of phenotype and genotype data. CA and BW performed phenotype harmonization. CA performed the statistical analysis. FH consulted on the statistical analysis. CA wrote the first draft of the manuscript. All authors contributed to manuscript revision, read, and approved the submitted version.

## 7 Acknowledgments

We wish to thank all of the participants in the Vitamin Intervention for Stroke Prevention trial that made this study possible. The authors acknowledge Research Computing at The University of Virginia for providing computational resources and technical support that have contributed to the results reported within this publication. URL: https://rc.virginia.edu

## 8 Funding

The GWAS component of the VISP study was supported by the United States National Human Genome Research Institute (NHGRI), Grant U01 HG005160 (PI Michèle Sale & Bradford Worrall), as part of the Genomics and Randomized Trials Network (GARNET). Genotyping services were provided by the Johns Hopkins University Center for Inherited Disease Research (CIDR), which is fully funded through a federal contract from the NIH to the Johns Hopkins University. Assistance with data cleaning was provided by the GARNET Coordinating Center (U01 HG005157; PI Bruce S Weir). Study recruitment and collection of datasets for the VISP clinical trial were supported by an investigator-initiated research grant (R01 NS34447; PI James Toole) from the United States Public Health Service, NINDS, Bethesda, Maryland. Control data for comparison with European ancestry VISP stroke cases were obtained through the database of genotypes and phenotypes (dbGAP) High Density SNP Association Analysis of Melanoma: Case-Control and Outcomes Investigation (phs000187.v1.p1; R01CA100264, 3P50CA093459, 5P50CA097007, 5R01ES011740, 5R01CA133996, HHSN268200782096C; PIs Christopher Amos, Qingyi Wei, Jeffrey E. Lee). For VISP stroke cases of African ancestry, a subset of the Healthy Aging in Neighborhoods of Diversity across the Life Span study (HANDLS) were used as stroke free controls. HANDLS is funded by the National Institute of Aging (1Z01AG000513; PI Michele K. Evans).

## 9 Disclosures

The authors declare that the research was conducted in the absence of any commercial or financial relationships that could be construed as a potential conflict of interest. Dr. Bradford Worrall is the Deputy Editor for the journal *Neurology*.

## 10 Data Availability Statement

The genetic associations results generated by this study can be found in the Cerebrovascular Disease Knowledge Portal (https://cd.hugeamp.org/XXXX).

## Notes

### Competing Interest Statement

The authors have declared no competing interest.

### Author Declarations

This study was approved by the internal review boards (IRBs) of Wake Forest University School of Medicine, University of North Carolina at Chapel Hill School of Medicine as well as individual recruiting sites in accordance with the declaration of Helsinki. All patients provided written informed consent [12]. However, ten sites did not approve the genetic portion of the study resulting in 2,100 genotyped patients.

